# Evaluating the Usability of Co-Created Public-Facing Knowledge Mobilization Resources for the Canadian Guidelines for Post COVID-19 Condition

**DOI:** 10.64898/2026.01.29.26344622

**Authors:** SA Elliott, S Cyrkot, SD Scott, H Travis, A Motilall, W Wiercioch, JP Pardo, HJ Schünemann, R Nieuwlaat, L Hartling

## Abstract

**Background:** The Canadian Guidelines for Post COVID-19 Condition (CAN-PCC), along with related knowledge mobilization (KM) resources, have been developed to support key interest-holders (e.g., public, healthcare providers, and policymakers) to make informed health decisions related to post COVID-19 condition (PCC). The purpose of this report is to describe the process and findings of our usability testing with members of the public on eight public-facing KM resources to support the CAN-PCC guidelines. These KM resources consist of an online resource sheet and an interactive infographic on four of the PCC guideline topics (1. prevention of PCC; 2. testing, identification and diagnosis; 3. pharmacological and non-pharmacological interventions; and 4. pediatric and adolescent health) that have been co-developed with interest-holders to enhance accessibility and understanding of the recommendations.

**Methods:** A convenience sample of members of the public were recruited online through our organizations’ pre-established networks. Each participant was randomly assigned to review one of eight resources before completing a questionnaire, developed by the research team and pilot-tested by a national Public Advisory Group that was formed to inform this work. The questionnaire contained 24 usability questions (5-point Likert scale, multiple response questions, open-text) that assessed: (1) purpose and organization, (2) credibility, (3) usefulness, (4) desirability, (5) value, and (6) accessibility. Demographic information was also collected. Descriptive statistics and measures of central tendency were calculated to summarize usability and demographic data. Independent t-tests were used to evaluate differences in mean scores between resource formats for each guideline topic. Open-ended responses were analyzed using thematic content analysis.

**Results:** We completed usability testing with 64 participants (n=8 assessments per resource) between March and April 2025. All eight resources were highly rated with average mean scores ranging from 3.8 to 4.6 on the 5-point scale, where five indicated ‘Strongly Agree’. No statistically significant differences were observed between total scores for the different resources, however, differences across some domains (e.g., credibility, desirability, value, accessibility; p<0.05) suggested that each resource type offers distinct strengths depending on the feature being assessed. Strengths of the resources included ease of understanding, clear and effective visual designs, and an empowering tone (e.g., sharing practice tips). Areas for improvement included clarifying the overall purpose and content related to the recommendations, as well as enhancing the visibility of embedded links within the resources.

**Conclusions:** Usability testing provided valuable insights into the usefulness and clarity of the resources. Final resources were made publicly available on the CAN-PCC website (https://canpcc.ca/resources/#tab-content-patients-and-general-public) in April 2025. We further disseminated the resources through social media channels and other pre-established networks. The findings will guide future guideline-related KM product development to ensure resources are useful, accessible, user-friendly and better meet the needs of the public.

## Introduction

Canadian Guidelines for Post COVID-19 Condition (CAN-PCC) were developed to support Canadians to make informed health decisions on the long-term effects experienced after a COVID-19 infection^(1)^. As of March 2025, the guidelines outline over 100 recommendations and good practice statements across six key guideline topics (GT): (GT1) Prevention of Post COVID-19 Condition (PCC); (GT2) Testing, Identification and Diagnosis related to PCC; (GT3) Pharmacological and Non-Pharmacological Interventions on PCC; (GT4) Neurological and Psychiatric topics; (GT5) Pediatric and Adolescent topics on PCC; and (GT6) Health Care Services and Systems, Social Support. The guidelines aim to provide clear, evidence-based recommendations to improve patient care and public health responses to PCC^(2)^.

As part of this initiative, the CAN-PCC Collaborative engaged teams across Canada to develop knowledge mobilization (KM) resources (e.g., webinars, social media blog posts, interactive infographics) to ensure that its guidelines for PCC are accessible and effectively implemented across diverse communities. These resources aim to bridge gaps between research, healthcare providers, policymakers, and the public by translating evidence-based recommendations into practical tools^(3)^.

Following the drafting of recommendations for each GT, we collaboratively co-developed eight public-facing KM resources in partnership with key interest-holders^(4)^, including a national Public Advisory Group which was developed to inform this work (eight members from across Canada with and without lived PCC experiences), and the CAN-PCC Collaborative (Guideline Teams, KM Committee, and Equity Oversight Committee) between August 2024 and April 2025^(5)^. These resources were also informed by lived experiences and information needs identified through focus groups and interviews with the general public^(6)^. They were designed to enhance the accessibility, clarity, and usability of the recommendations and good practice statements (released prior to April 2025) covering four CAN-PCC GT (GT1: 22 recommendations, 2 good practice statements; GT2: 16 recommendations, 3 good practice statements; GT3: 19 recommendations 3 good practice statements; GT5: 15 recommendations, 5 good practice statements). These four topics were selected as they are relevant to the general public, including parents and caregivers. Our iterative design process integrated feedback from a range of perspectives to ensure the resources were relevant, clear, and user-friendly. For each topic, one online resource sheet (ORS) and one interactive infographic were created and refined. Following the development of these eight resources, we conducted usability testing to ensure the final materials were acceptable, engaging, and useful^(7)^. After refinement based on usability testing results, the eight KM resources were made publicly available in April 2025 on the CAN-PCC website (https://canpcc.ca), and shared with relevant interest-holders (e.g., via social media, organization email list servs). The ORS are available to view in eight different languages (English, French, Arabic, Chinese, Punjabi, Spanish, Tagalog, Ukrainian) and the interactive infographics are available to view in two different languages (English, French).

This report outlines the process and findings of our usability testing, which guided the final development of KM resources.

## Methods

### Overview, Sampling and Recruitment

Usability testing was one component of a broader cross-sectional descriptive study designed to explore the experiences and information needs of individuals living with PCC, while also informing the co-development of the CAN-PCC KM resources using an iterative design process^(6)^. The University of Alberta Health Research Ethics Board (Pro00140055) and the Hamilton Integrated Research Ethics Board (Pro16317) approved the research study and participant consent was obtained prior to any data collection.

Members of the public (with or without PCC, including caregivers) were eligible for study enrollment to complete usability testing if they identified as an adult over 18 years, lived in Canada, were able to understand English, and had an electronic device with access to the Internet (including email). Members of the public were recruited online (e.g., email lists, e-newsletters, word of mouth) through a convenience sample gathered from pre-established networks and connections (e.g., University of Alberta, CAN-PCC,

Public Advisory Group) in March and April 2025. After study completion, participants were compensated with one CAD $10 electronic gift card for their time.

### Study Components

#### Usability Testing

Interested participants who viewed the recruitment materials contacted the research team by email and were sent a link to complete a usability questionnaire (5-10 minutes) on one of the eight KM resources (**Supplemental 1**) which were allocated using 1:1 randomization. All participants had an opportunity to review the study information letter and ask questions before providing informed consent by completing the questionnaire. The questionnaire consisted of 24 questions assessing six domains: (1) purpose and organizations, (2) credibility, (3) usefulness, (4) desirability, (5) value, and (6) accessibility, along with nine demographic questions (e.g., age, gender, education, ethnicity). The question responses were a combination of a 5-point Likert scale, multiple response questions, and open-text. The questionnaire was developed by the research team based on previously published usability questions and structured following the Morville’s Honeycomb model of user experience (**Supplemental 2**)^(8-12)^. The questionnaire was refined through pilot-testing with our Public Advisory Group. Data were collected and managed using REDCap (Research Electronic Data Capture), hosted at the University of Alberta^(13, 14^).

#### Data analysis

Usability data were cleaned and analyzed using Microsoft Excel (2019, Microsoft Corporation). Descriptive statistics and measures of central tendency were calculated for demographic and Likert-scale responses, with scores ranging from 5 (Strongly Agree) to 1 (Strongly Disagree). Data was checked for normality using the Shapiro-Wilk test. Independent t-tests were used to evaluate differences in mean scores between resource formats (ORS versus interactive infographic) for each GT. Open-ended responses were thematically analyzed using an inductive approach^(15)^, with researchers reviewing and discussing comments to inform potential revisions to the eight KM resources.

#### Sample Size

Research has shown that approximately 80% of usability issues can be identified with as few as four to five participants. Importantly, the most severe problems tend to emerge early in the testing process, while additional participants contribute diminishing returns in terms of new insights ^(16)^. Based on this evidence, a sample size of eight participants per tool was chosen to exceed the minimum threshold, ensuring both common and less frequent issues were captured efficiently.

## Results

We completed usability testing with 64 participants (n=8 assessments per resource) between March and April 2025. Demographic characteristics are presented in **Table 1**. Approximately 40% of participants (n=26) reported having lived PCC experience and/or were caregivers of a person with lived PCC. In contrast, 52% (n=33) did not and 8% (n=5) did not disclose their status. Most participants were from Alberta (70%, n=45), followed by Ontario (16%, n=10), and British Columbia (9%, n=6). Education levels ranged from high school diplomas (3%, n=2) to post-secondary or graduate degrees (86%, n=55). Participants most commonly identified as White (50%, n=32), East Asian (14%, n=9), or South Asian (14%, n=9).

**Table 1.**
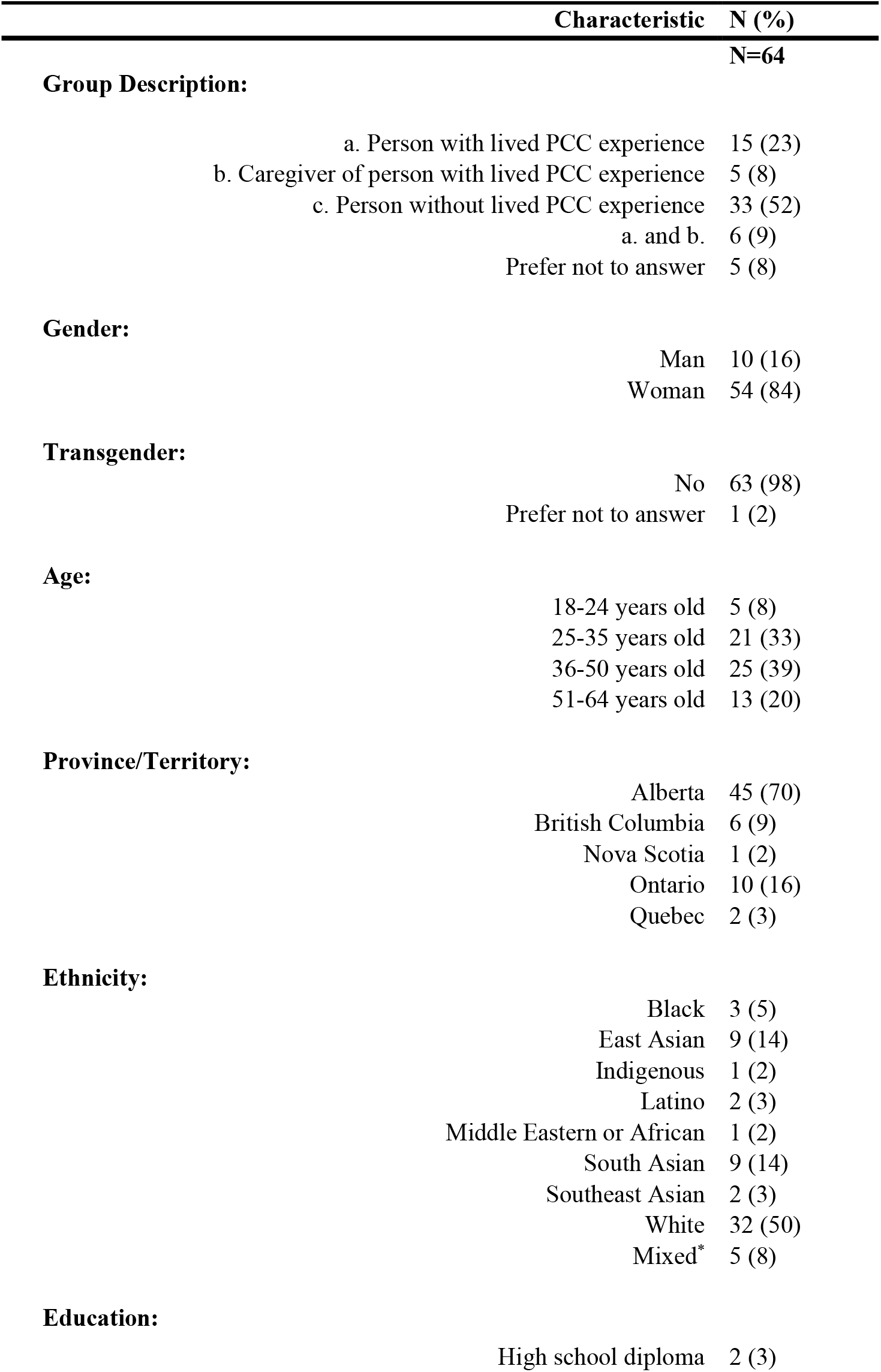
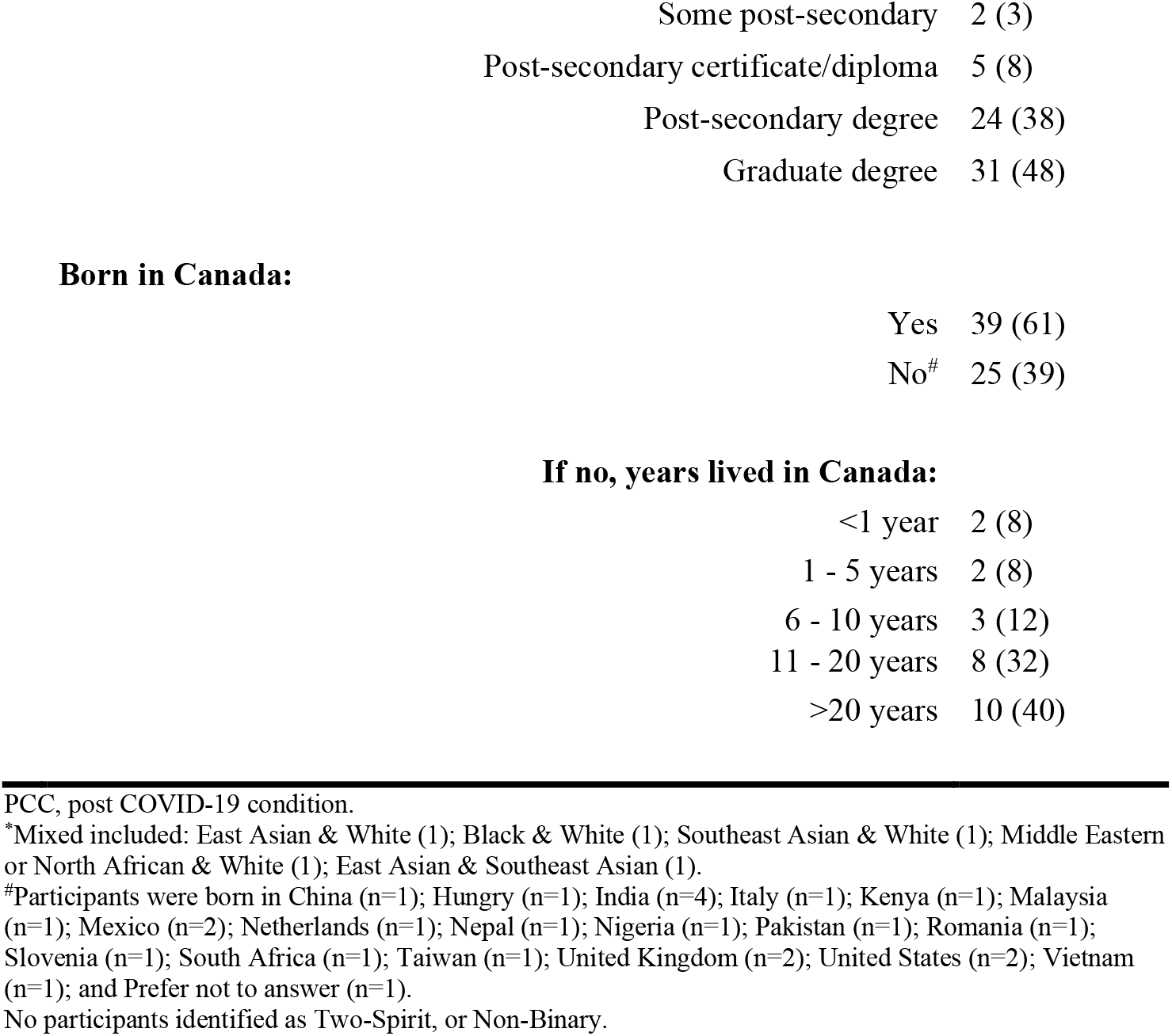
Demographic characteristics of participants who assessed the usability of the eight knowledge mobilization resources on post COVID-19 condition.

| Characteristic |  | N (%) |
| --- | --- | --- |
| **Group Description:** |  | **N=64** |
| a. Person with lived PCC experience |  | 15 (23) |
| b. Caregiver of person with lived PCC experience |  | 5 (8) |
| c. Person without lived PCC experience |  | 33 (52) |
| a. and b. |  | 6 (9) |
| Prefer not to answer |  | 5 (8) |
| **Gender:** |  |  |
| Man |  | 10 (16) |
| Woman |  | 54 (84) |
| **Transgender:** |  |  |
| No |  | 63 (98) |
| Prefer not to answer |  | 1 (2) |
| **Age:** |  |  |
| 18-24 years old |  | 5 (8) |
| 25-35 years old |  | 21 (33) |
| 36-50 years old |  | 25 (39) |
| 51-64 years old |  | 13 (20) |
| **Province/Territory:** |  |  |
| Alberta |  | 45 (70) |
| British Columbia |  | 6 (9) |
| Nova Scotia |  | 1 (2) |
| Ontario |  | 10 (16) |
| Quebec |  | 2 (3) |
| **Ethnicity:** |  |  |
| Black |  | 3 (5) |
| East Asian |  | 9 (14) |
| Indigenous |  | 1 (2) |
| Latino |  | 2 (3) |
| Middle Eastern or African |  | 1 (2) |
| South Asian |  | 9 (14) |
| Southeast Asian |  | 2 (3) |
| White |  | 32 (50) |
| Mixed* |  | 5 (8) |
| **Education:** |  |  |
| High school diploma |  | 2 (3) |

|  |  |
| --- | --- |
| Some post-secondary | 2 (3) |
| Post-secondary certificate/diploma | 5 (8) |
| Post-secondary degree | 24 (38) |
| Graduate degree | 31 (48) |

**Born in Canada:**
|  |  |
| --- | --- |
| Yes | 39 (61) |
| No<sup>#</sup> | 25 (39) |

**If no, years lived in Canada:**
|  |  |
| --- | --- |
| <1 year | 2 (8) |
| 1 - 5 years | 2 (8) |
| 6 - 10 years | 3 (12) |
| 11 - 20 years | 8 (32) |
| >20 years | 10 (40) |

PCC, post COVID-19 condition.
\*Mixed included: East Asian & White (1); Black & White (1); Southeast Asian & White (1); Middle Eastern or North African & White (1); East Asian & Southeast Asian (1).
<sup>#</sup>Participants were born in China (n=1); Hungary (n=1); India (n=4); Italy (n=1); Kenya (n=1); Malaysia (n=1); Mexico (n=2); Netherlands (n=1); Nepal (n=1); Nigeria (n=1); Pakistan (n=1); Romania (n=1); Slovenia (n=1); South Africa (n=1); Taiwan (n=1); United Kingdom (n=2); United States (n=2); Vietnam (n=1); and Prefer not to answer (n=1).
No participants identified as Two-Spirit, or Non-Binary.

All eight KM resources were highly rated. The average total scores for ORS and interactive infographics across the topics ranged from 4.1 to 4.2 and 3.8 to 4.6, respectively, on a 5-point scale, with five representing ‘Strongly Agree’. Mean scores by GT and domain are presented in **Table 2** for the ORS and **Table 3** for the interactive infographics. When comparing how participants rated the ORS and interactive infographic resources across several domains, the interactive infographics were seen as more valuable for GT1 (mean score: ORS 3.4 vs. infographic 4.4, p=0.03), more desirable to use for GT2 (mean score: ORS 4.0 vs. infographic 4.7, p=0.01), and easier to access for GT3 (mean score: ORS 3.9 vs. infographic 4.7, p=0.04). Conversely, participants rated the GT5 ORS as more credible than the interactive infographic (mean score: ORS 4.5 vs. infographic 3.4, p=0.02). No statistically significant differences were found in other areas such as overall usefulness, organization, or total scores. These results suggest that while both resource types were generally well received, each had its own strengths depending on what aspect was being evaluated.

**Table 2.**
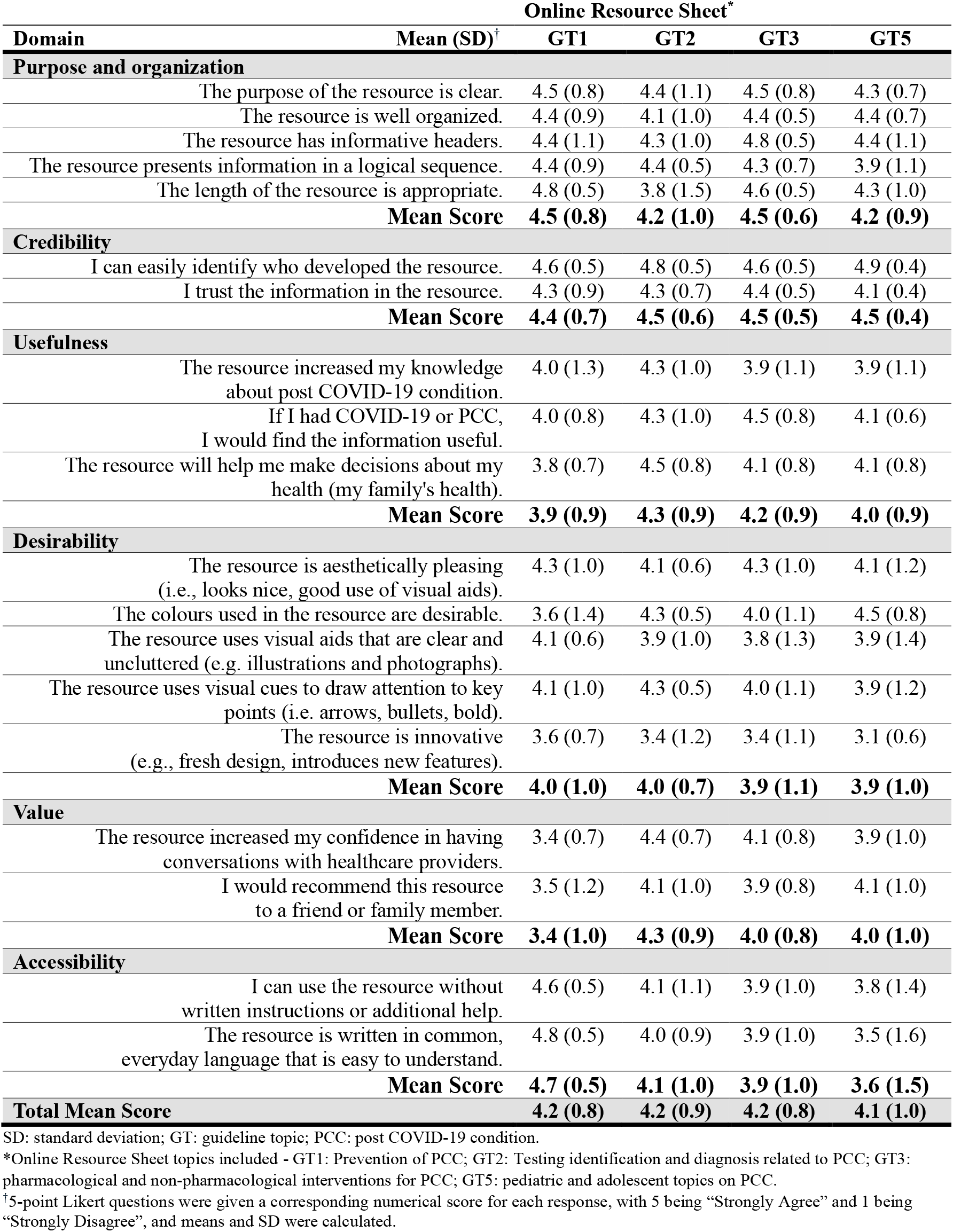
Means (SD) of participant responses to the usability questionnaire evaluating the four online resource sheets for post COVID-19 condition.

**Table 3.** Means (SD) of participant responses to the usability questionnaire evaluating the four interactive infographics for post COVID-19 condition.

|  |  | Interactive Infographic* |  |  |  |
| --- | --- | --- | --- | --- | --- |
| Domain | Mean (SD)<sup>†</sup> | GT1 | GT2 | GT3 | GT5 |
| Purpose and organization |  |  |  |  |  |
| The purpose of the resource is clear. |  | 4.5 (0.5) | 4.6 (0.5) | 4.6 (0.5) | 4.0 (1.4) |
| The resource is well organized. |  | 4.0 (0.8) | 4.6 (0.5) | 4.6 (0.5) | 3.9 (1.1) |
| The resource has informative headers. |  | 4.3 (0.7) | 4.8 (0.5) | 4.9 (0.4) | 4.1 (1.1) |
| The resource presents information in a logical sequence. |  | 4.1 (0.8) | 4.5 (0.5) | 4.3 (1.2) | 4.3 (1.0) |
| The length of the resource is appropriate. |  | 4.1 (1.0) | 3.6 (0.7) | 4.3 (1.0) | 3.9 (1.4) |
|  | Mean Score | 4.2 (0.8) | 4.4 (0.6) | 4.5 (0.7) | 4.0 (1.2) |
| Credibility |  |  |  |  |  |
| I can easily identify who developed the resource. |  | 4.1 (1.0) | 4.5 (0.5) | 4.0 (1.2) | 3.6 (1.4) |
| I trust the information in the resource. |  | 4.0 (0.8) | 4.8 (0.5) | 4.0 (0.8) | 3.1 (1.6) |
|  | Mean Score | 4.1 (0.9) | 4.6 (0.5) | 4.0 (1.0) | 3.4 (1.5) |
| Usefulness |  |  |  |  |  |
| The resource increased my knowledge about post COVID-19 condition. |  | 3.9 (1.2) | 4.8 (0.5) | 3.8 (1.8) | 3.0 (1.1) |
| If I had COVID-19 or PCC, I would find the information useful. |  | 4.0 (0.8) | 4.5 (1.1) | 4.5 (0.5) | 3.3 (1.4) |
| The resource will help me make decisions about my health (my family's health). |  | 3.9 (1.0) | 4.4 (0.7) | 4.4 (0.9) | 3.3 (1.4) |
|  | Mean Score | 3.9 (1.0) | 4.5 (0.8) | 4.2 (1.1) | 3.2 (1.3) |
| Desirability |  |  |  |  |  |
| The resource is aesthetically pleasing (i.e., looks nice, good use of visual aids). |  | 4.3 (0.7) | 4.8 (0.5) | 4.6 (0.7) | 4.3 (1.4) |
| The colours used in the resource are desirable. |  | 4.5 (0.5) | 4.9 (0.4) | 4.5 (0.9) | 4.1 (1.0) |
| The resource uses visual aids that are clear and uncluttered (e.g. illustrations and photographs). |  | 4.4 (0.7) | 4.6 (0.5) | 4.8 (0.5) | 4.4 (0.5) |
| The resource uses visual cues to draw attention to key points (i.e. arrows, bullets, bold). |  | 4.1 (1.0) | 4.6 (0.5) | 4.8 (0.5) | 4.4 (0.5) |
| The resource is innovative (e.g., fresh design, introduces new features). |  | 4.4 (0.5) | 4.5 (0.8) | 4.3 (1.2) | 3.5 (1.3) |
|  | Mean Score | 4.3 (0.7) | 4.7 (0.5) | 4.6 (0.8) | 4.1 (0.9) |
| Value |  |  |  |  |  |
| The resource increased my confidence in having conversations with healthcare providers. |  | 4.3 (0.9) | 4.5 (0.5) | 4.0 (1.1) | 3.1 (1.4) |
| I would recommend this resource to a friend or family member. |  | 4.5 (0.8) | 4.9 (0.4) | 4.4 (0.7) | 2.9 (1.6) |
|  | Mean Score | 4.4 (0.8) | 4.7 (0.4) | 4.2 (0.9) | 3.0 (1.5) |
| Accessibility |  |  |  |  |  |
| I can use the resource without written instructions or additional help. |  | 4.6 (0.5) | 4.8 (0.5) | 4.6 (0.5) | 4.4 (1.1) |
| The resource is written in common, everyday language that is easy to understand. |  | 4.5 (0.5) | 4.8 (0.5) | 4.8 (0.5) | 4.5 (0.8) |
|  | Mean Score | 4.6 (0.5) | 4.8 (0.5) | 4.7 (0.5) | 4.4 (0.9) |
| Total Mean Score |  | 4.2 (0.8) | 4.6 (0.6) | 4.4 (0.8) | 3.8 (1.2) |

SD: standard deviation; GT: guideline topic; PCC: post COVID-19 condition.
\*Interactive infographic topics included - GT1: Prevention of PCC; GT2: Testing identification and diagnosis related to PCC; GT3: pharmacological and non-pharmacological interventions for PCC; GT5: pediatric and adolescent topics on PCC.
<sup>†</sup>5-point Likert questions were given a corresponding numerical score for each response, with 5 being “Strongly Agree” and 1 being “Strongly Disagree”, and means and SD were calculated.

**Table 4a.** Strengths and areas for improvement identified by participants regarding the four online resource sheets.

|  | Description | Illustrative Comment |
| --- | --- | --- |
| **Strengths** |  |  |
| Appropriate length with a clear and effective visual design | Concise and quick to read. The layout, use of icons, and color scheme contributes to a professional presentation. | *"It is straight to the point and provides essential information."* (GT2, Participant 02)<br>*"Provides additional direction to garner more information, if desired."* (GT1, Participant 03) |
| Easy to understand with links for more information | Reads well and in plain language, with links to find more information, if desired. | *"I like that it's easy to read and understand."* (GT1, Participant 05)<br>*"The links provided for further information about PCC were well laid out and easy to navigate."* (GT5, Participant 06) |
| Empowering | Provides informative and educational information. Offers practical tips and next steps (e.g., talk to a healthcare provider for more information). | *"I like that it provides information about the tests that could be used to test for PCC which I think can be really empowering to people to have better informed conversations with their physician"* (GT2, Participant 05) |
| **Area for improvement** |  |  |
| Clarify purpose and messaging | Make the purpose and key messages of the resource clear and easy to understand. | *"I felt like parts of it were confusing in terms of what I was supposed to gather from certain sections."* (GT2, Participant 05) |
| Address content depth, refine language and text volume, and ensure embedded links to additional content are visible and accessible | Find a balance between including key details while also limiting word-count, using clear language, explaining complex terms (e.g., certainty of evidence), and improving the visibility of links to find additional information. | *"I desired more information after reading it."* (GT1, Participant 01)<br>*"The fine print was confusing regarding the certainty of evidence. I was unsure if that means the medications and supplements listed are good or bad."* (GT3, Participant 04) |

GT, guideline topic; GT1: Prevention of PCC; GT2: Testing identification and diagnosis related to PCC; GT3: pharmacological and non-pharmacological interventions for PCC; GT5: pediatric and adolescent topics on PCC.

**Table 4b.**
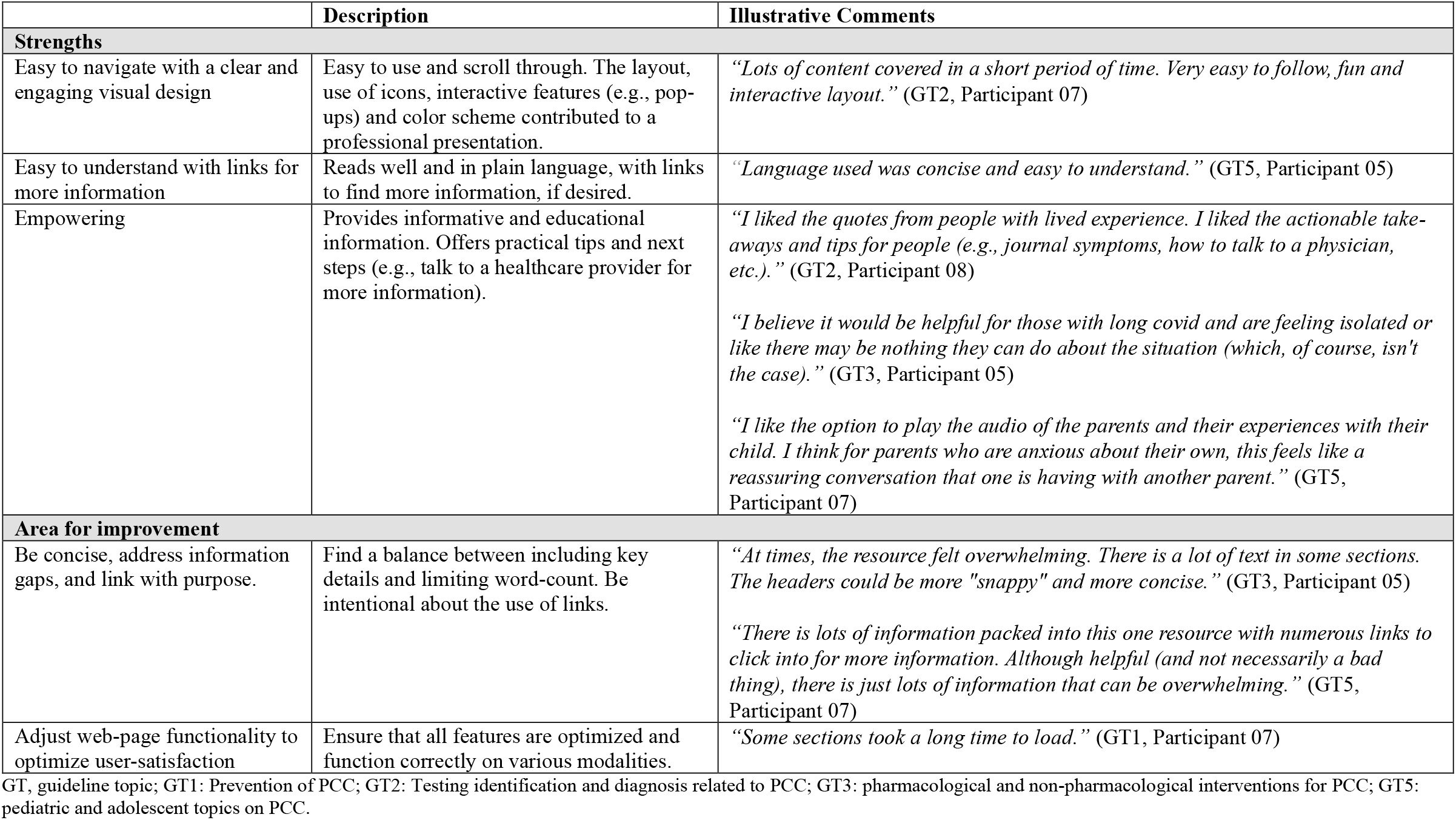
Strengths and areas for improvement identified by participants regarding the four interactive infographics.

Open-ended comments that addressed resource strengths and areas for improvement, as well as factors influencing trust towards using the resources, are summarized in **Table 4** and **Table 5**. The resources were assessed as being concise, well-designed, and easy to understand, using clear language and effective visuals. Participants noted that the resources offered practical information and guidance for individuals seeking more knowledge about PCC. However, areas for improvement included clarifying the purpose and messaging, refining language for clarity, ensuring a balance between detail and readability, and enhancing the visibility of embedded links. Some readers found certain terms confusing (e.g., certainty of evidence, recommendation strength) and desired more in-depth information.

**Table 5.** Elements self-identified by participants that enhance trust towards using the resources.

| Elements | Description | Illustrative quote from participants |
| --- | --- | --- |
| Clearly listed and reputable information sources | Institutional names and logos were displayed on the resources to show who developed or funded it. Information sources that were more familiar to participants were generally perceived as more trustworthy. | *"I trust the information because the creators are clearly stated at the top of the resource, and there is a statement at the bottom of the resource citing financial contribution."* (Interactive Infographic, GT3, Participant 05)<br>*"The inclusion of a university and PHAC makes me trust the resource at 70%; however, the inclusion of organizations which are unfamiliar to me (e.g., Cochrane Canada) reduced my trust as I don't know who they are and what their reliability is."* (Online Resource Sheet, GT5, Participant 04) |
| Clear, informative, and professional content and design | Clearly presented (e.g., plain language), informative (e.g., appropriate level of detail), and professional (e.g., colour, layout) information was general perceived as more trustworthy. | *"The information which is on document is clear, easy to understand and straightforward."* (Online Resource Sheet, GT2, Participant 07)<br>*"The resource appears to be free of typo and looks official, which suggest that it is created from a credible source."* (Online Resource Sheet, GT3, Participant 04) |
| Content that resonated with lived experiences and prior knowledge | Information that aligned with lived experiences and prior knowledge were generally perceived as more trustworthy. | *"Because I know things about PCC and the things that are mentioned in the infographic align with my current knowledge."* (Interactive Infographic, GT1, Participant 05)<br>*"As some of the information contradicts my lived experience [low-dose naltrexone], I trust it less."* (Interactive Infographic, GT3, Participant 03) |

GT, guideline topic; GT1: Prevention of PCC; GT2: Testing identification and diagnosis related to PCC; GT3: pharmacological and non-pharmacological interventions for PCC; GT5: pediatric and adolescent topics on PCC.

## Discussion and Conclusions

Providing the general public with research evidence in user-friendly formats (e.g., interactive infographics, ORS) can increase knowledge and understanding of the CAN-PCC guidelines. This in turn can empower and support individuals to make informed decisions and participate in shared-decision making with their healthcare providers^(17)^. Additionally, increasing ‘accessibility’ to evidence (through increasing awareness and knowledge) in lay formats, and addressing the target audience’s specific information needs, will have the potential to increase use of the recommendations (i.e., if people are aware of the recommendations and can understand them, they will be more likely to use them).

We developed and tested eight KM resources to facilitate the uptake and dissemination of 72 recommendations and 13 good practice statements from four Canadian CAN-PCC guidelines nationwide. These resources were designed to ensure that the guidelines are accessible and easily understood by the general public.

Mean total scores for the interactive infographics exhibited a wider range compared to the ORS (despite not being statistically significant), potentially reflecting varied responses to the narrative framing of certain topics (e.g., GT5, Pediatrics) which may have elicited more diverse reactions. Differences between individuals with lived experiences and those without lived experiences (including caregivers) were not explored, however; future research could benefit from examining these distinctions.

The ORS were visually clear and professionally designed, with accessible language that made them easy to understand and contained content that offered practical next steps. Their main limitation was that the purpose and key messages of some recommendations were not always explicit, requiring clearer articulation to strengthen their impact. In contrast, the interactive infographics stood out for their engaging design and ease of navigation, supported by interactive features, accessible links, and practical guidance. However, they required refinement in streamlining text, using links more intentionally, and ensuring that webpage functionality operated smoothly across different modalities to maximize user satisfaction.

Trust in the resources was influenced by several factors. The inclusion of reputable information sources, such as institutional names and logos, increased credibility, although unfamiliar organizations sometimes led to skepticism. Clear, informative, and professionally presented content using plain language, an appropriate level of detail, and a polished design enhanced perception of reliability. Additionally, alignment with lived experiences and prior knowledge played a crucial role; participants found information that matched their personal understanding of PCC to be more trustworthy, whereas contradictions with their experiences led to doubt. Ensuring clarity, accessibility, and resonance with users’ backgrounds can significantly strengthen the perceived credibility of knowledge mobilization efforts.

A study strength included achieving our targeted sample size which supported a robust and reliable testing process for useability. However, we acknowledge that recruitment was not conducted across all regions of Canada due to feasibility and time constraints, thus limiting generalizability. Future studies would benefit from having time and resources available to develop a more comprehensive recruitment strategy to increase recruitment at a national level to better capture regional differences (despite major similarities within the healthcare systems). It may also be beneficial to amend the inclusion criteria to solely recruit parent participants to review GT5 (Pediatrics) given their lived experiences caring for a child, as not all participants who completed usability testing on GT5 identified as a parent of a child under 18 years. Despite conducting usability testing on the English resources, testing was not conducted on all linguistically translated resources (e.g., Arabic, Spanish). Future research warrants usability testing on translated resources as certain nuances in languages as well as culture differences may impact the acceptance and adoption of the resources among diverse populations.

Evaluation results indicate that actively involving groups with lived experience in the development of KM resources is an effective approach for creating highly usable and widely accepted resources. These resources are now available as part of a broader collection of CAN-PCC resources developed to support individuals with PCC, healthcare providers, and policy makers.

## Acknowledgements

We would like to thank the members of our Public Advisory Group for supporting this work and pilot testing the usability questionnaire.

## Data Availability

All data produced in the present study are available upon reasonable request to the authors.

## Abbreviations

CAN-PCC: Canadian Guidelines for Post COVID-19 Condition
GT: Guideline topic
KM: Knowledge mobilization
ORS: Online Resource Sheet
PCC: Post COVID-19 Condition
REDCap: Research Electronic Data Capture

---

**Supplemental 1A:**
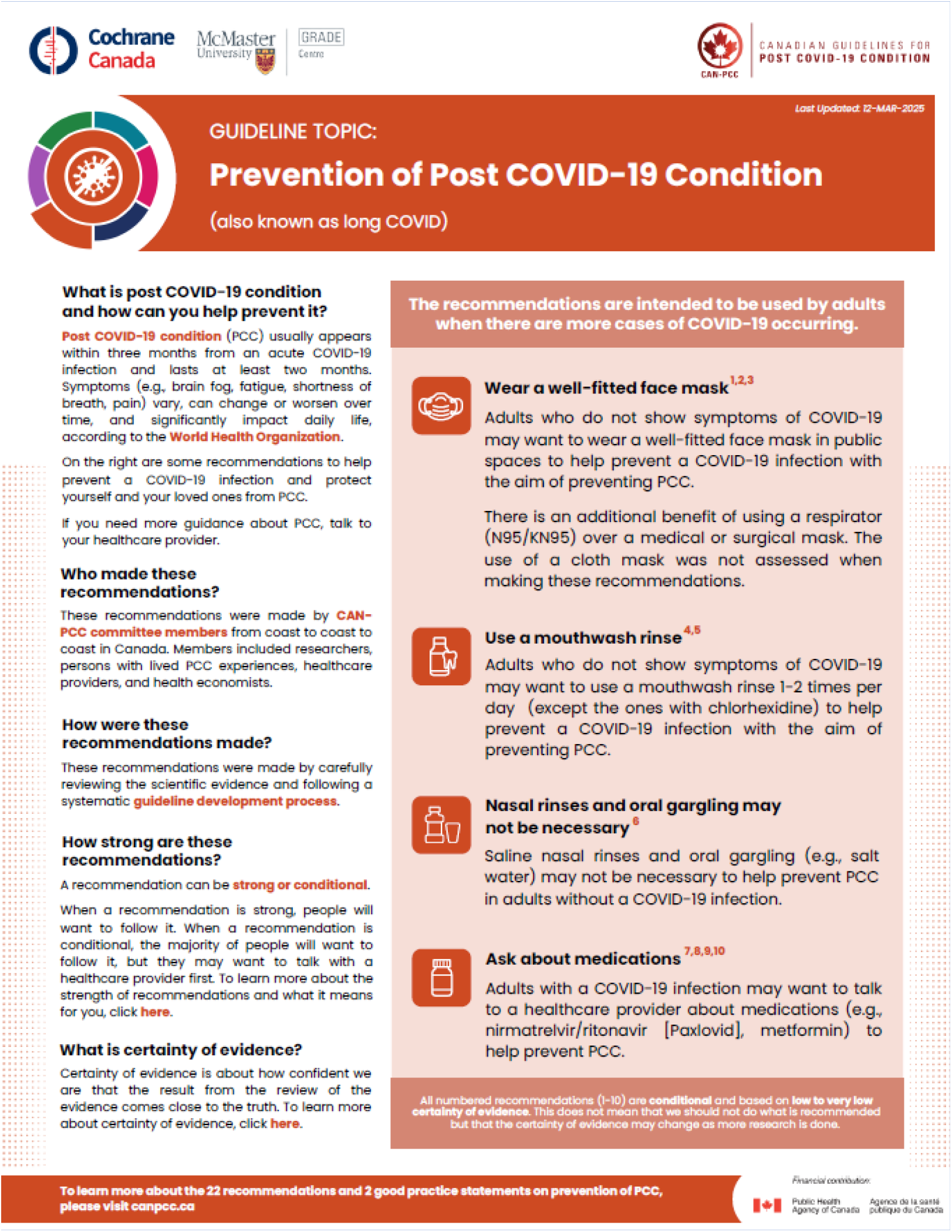
Online Resource Sheet Screenshots. Example of GT1: Prevention of Post COVID-19 Condition

**Supplemental 1B:**
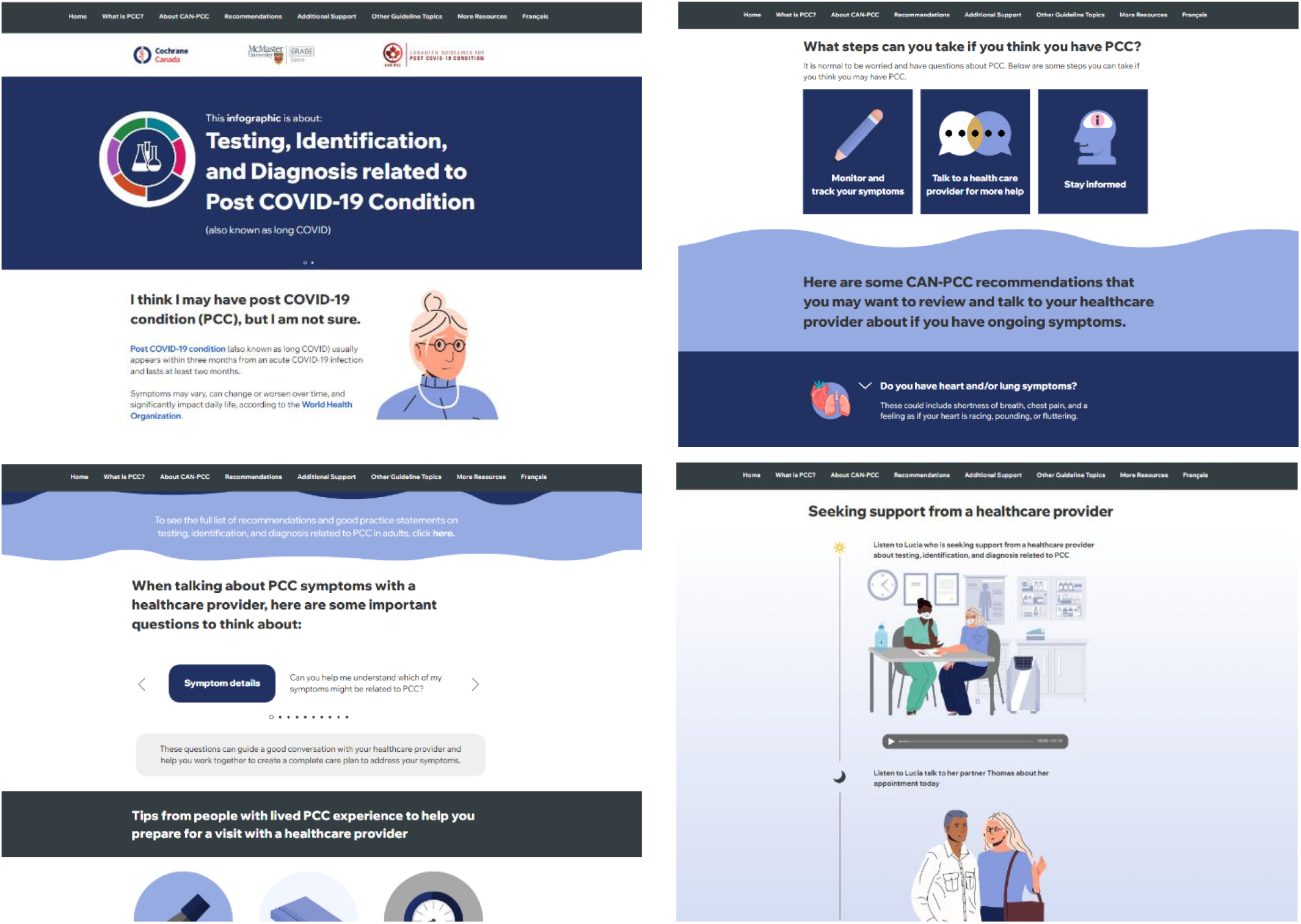
Interactive Infographic Screenshots. Example of GT2: Testing, Identification and Diagnosis Related to Post COVID-19 Condition

## Supplemental 2: Usability Questionnaire

### SECTION 1: PURPOSE AND ORGANIZATION

#### 1. The purpose of the resource is clear

a. Strongly agree
b. Agree
c. Neither agree nor disagree
d. Disagree
e. Strongly disagree

#### 2. The resource is well organized

a. Strongly agree
b. Agree
c. Neither agree nor disagree
d. Disagree
e. Strongly disagree

#### 3. The resource has informative headers

a. Strongly agree
b. Agree
c. Neither agree nor disagree
d. Disagree
e. Strongly disagree

#### 4. The resource presents information in a logical sequence

a. Strongly agree
b. Agree
c. Neither agree nor disagree
d. Disagree
e. Strongly disagree

#### 5. The length of the resource is appropriate

a. Strongly agree
b. Agree
c. Neither agree nor disagree
d. Disagree
e. Strongly disagree

### SECTION 2: CREDIBILITY

#### 6. I can easily identify who developed the resource

a. Strongly agree
b. Agree
c. Neither agree nor disagree
d. Disagree
e. Strongly disagree

#### 7. I trust the information in the resource

a. Strongly agree
b. Agree
c. Neither agree nor disagree
d. Disagree
e. Strongly disagree

#### 7a. What is it that leads you or does not lead you to believe that?

### SECTION 3: USEFULNESS

#### 8. The resource increased my knowledge about post COVID-19 condition

a. Strongly agree
b. Agree
c. Neither agree nor disagree
d. Disagree
e. Strongly disagree

#### 9. What do you think about the information in this resource? *(check all that apply)*

a. This information teaches me something new
b. This information allows me to validate what I do or I did
c. This information reassures me
d. This information refreshes my memory
e. This information motivates me to learn more
f. I think there is a problem with this information
g. I disagree with this information

#### 10. If I had COVID-19 or post COVID-19 condition, I would find the information useful

a. Strongly agree
b. Agree
c. Neither agree nor disagree
d. Disagree
e. Strongly disagree

#### 11. How would you use this information? *(check all that apply)*

a. This information would help me to better understand something
b. If I did not know what to do, this information would help me to do something
c. If I knew what to do, this information would convince me to do it
d. I would use this information to do something in a different manner
e. I would use this information to speak with someone (e.g. doctor)
f. I do not see myself using this information
g. Other

*(branched to 11, g. Other)*

#### 11a. Other (please specify)

#### 12. The resource will help me make decisions about my health (my family’s health)

a. Strongly agree
b. Agree
c. Neither agree nor disagree
d. Disagree
e. Strongly disagree

### SECTION 4: DESIRABILTIY

#### 13. The resource is aesthetically pleasing (i.e. looks nice, good use of visual aids)

a. Strongly agree
b. Agree
c. Neither agree nor disagree
d. Disagree
e. Strongly disagree

#### 14. The colours used in the resource are desirable

a. Strongly agree
b. Agree
c. Neither agree nor disagree
d. Disagree
e. Strongly disagree

#### 15. The resource uses visual aids that are clear and uncluttered (e.g. illustrations and photographs)

a. Strongly agree
b. Agree
c. Neither agree nor disagree
d. Disagree
e. Strongly disagree

#### 16. The resource uses visual cues to draw attention to key points (i.e. arrows, bullets, bold)

a. Strongly agree
b. Agree
c. Neither agree nor disagree
d. Disagree
e. Strongly disagree

#### 17. The resource is innovative (e.g. fresh design, introduces new features)

a. Strongly agree
b. Agree
c. Neither agree nor disagree
d. Disagree
e. Strongly disagree

### SECTION 5: VALUE

#### 18. Do you expect any benefit from you using this information? *(check all that apply)*

a. This information will help me to improve my health or well-being
b. This information will help me to be less worried
c. This information will help me to prevent a problem or the worsening of a problem
d. This information will help me to handle a problem
e. This information will help me to be better prepared to speak with someone (e.g. doctor)
f. This information will help me to be more confident to make a decision about something with someone
g. No, I expect no benefits

#### 19. The resource increased my confidence if having conversations with a healthcare provider

a. Strongly agree
b. Agree
c. Neither agree nor disagree
d. Disagree
e. Strongly disagree

#### 20. I would recommend this resource to a friend or family member

a. Strongly agree
b. Agree
c. Neither agree nor disagree
d. Disagree
e. Strongly disagree

### SECTION 6: ACCESSIBILITY

#### 21. I can use the resource without written instructions or additional help

a. Strongly agree
b. Agree
c. Neither agree nor disagree
d. Disagree
e. Strongly disagree

#### 22. The resource is written in common, everyday language that is easy to understand

a. Strongly agree
b. Agree
c. Neither agree nor disagree
d. Disagree
e. Strongly disagree

#### 23. What did you like the most about the CAN-PCC resource?

#### 24. What did you like the least about the CAN-PCC resource?

### SECTION 7: DEMOGRAPHICS

#### 25. Which group description(s) apply best to you? (check all that apply)

*(post COVID-19 condition (also known as long COVID)*

a. Person with lived post COVID-19 condition experience
b. Caregiver for person with lived post COVID-19 condition experience (e.g. parent or guardian)
c. Person without lived post COVID-19 condition experience
d. Prefer not to answer

*(branched to 25)*

#### 25a. Please specify if there is additional information that you would like to share

#### 26. Which gender do you identify with most?

a. Male
b. Female
c. Non-binary
d. Gender fluid
e. Not sure or questioning
f. Another preferred term
g. Prefer not to answer

*(branched to 26)*

#### 26a. Another preferred term (please specify)

#### 27. Would you describe yourself as transgender?

a. Yes
b. No
c. Prefer not to answer

#### 28. Which of the following race and ethnicity groups best describes you? (check all that apply)

a. Black (i.e. African, Afro-Caribbean, African Canadian descent)
b. East Asian (i.e. Chinese, Korean, Japanese, Taiwanese descent)
c. Indigenous (i.e. First Nations, Métis, Inuk/Inuit descent)
d. Latino (i.e. Latin American, Hispanic descent)
e. Middle Eastern or North African (i.e. Arab, Persian, Afghan, Egyptian, Iranian, Lebanese, Turkish, other West Asian descent)
f. South Asian (i.e. East Indian, Pakistani, Bangladeshi, Sri Lankan, Indo-Caribbean, other South Asian descent)
g. Southeast Asian (i.e. Filipino, Vietnamese, Cambodian, Thai, Indonesia, other Southeast Asian descent)
h. White (i.e. European descent)
i. Not listed or other
j. Do not know
k. Prefer not to answer

*(branched to 28)*

#### 28a. Would you describe yourself as Two-Spirit?

a. Yes
b. No
c. Prefer not to answer

*(branched to 28)*

#### 28a Not listed or other (please specify)

#### 29. What is your age?

a. 18-24 years old
b. 25-35 years old
c. 36-50 years old
d. 51-64 years old
e. 65 years and older
f. Prefer not to answer

#### 30. What is your highest level of education?

a. Some high school
b. High school diploma
c. Some post-secondary
d. Post-secondary certificate/diploma
e. Post-secondary degree
f. Graduate degree
g. Other
h. Prefer not to answer

*(branched to 30)*

#### 30a. Other (please specify)

#### 31. Which province or territory do you currently live in?

a. Alberta
b. British Columbia
c. Manitoba
d. New Brunswick
e. Newfoundland and Labrador
f. Northwest Territories
g. Nova Scotia
h. Nunavut
i. Ontario
j. Prince Edward Island
k. Quebec
l. Saskatchewan
m. Yukon
n. Prefer not to answer

#### 32a. What is the primary language spoken in your home? (e.g. English) 32b. What other languages can you speak comfortably?

#### 33. Were you born in Canada?

a. Yes
b. No
c. Prefer not to answer

*(branched to 33)*

#### 33a. If you were not born in Canada, where were you born?

*(branched to 33)*

#### 33b. How long have you lived in Canada?

a. Less than 1 year
b. 1-5 years
c. 6-10 years
d. 11-20 years
e. More than 20 years

### END OF QUESTIONNAIRE

## References

1. Canadian Guidelines for Post COVID-19 Condition. Our Approach: McMaster University; 2025. Available from: https://canpcc.ca/our-approach/.

2. Canadian Guidelines for Post COVID-19 Condition. Guidelines: McMaster University; 2025. Available from: https://can-pcc.recmap.org/.

3. Straus SE, Tetroe J, Graham I. Defining knowledge translation. Cmaj. 2009;181(3-4):165–8.

4. Akl EA, Khabsa J, Petkovic J, Magwood O, Lytvyn L, Motilall A, et al. “Interest-holders”: A new term to replace “stakeholders” in the context of health research and policy. Cochrane Evidence Synthesis and Methods. 2024;2(11):e70007.

5. Canadian Guidelines for Post COVID-19 Condition. About Us: McMaster University; 2025. Available from: https://canpcc.ca/about-us/.

6. Elliott SA, Cyrkot S, Scott SD, Hartling L. Co-developing knowledge mobilization resources for long COVID: Insights into public experiences and information needs. In Development. 2025.

7. Keenan HL, Duke SL, Wharrad HJ, Doody GA, Patel RS. Usability: An introduction to and literature review of usability testing for educational resources in radiation oncology. Tech Innov Patient Support Radiat Oncol. 2022;24:67–72.

8. Morville P. User Experience Design 2004. Available from: http://semanticstudios.com/user_experience_design/.

9. Elliott SA, Scott SD, Charide R, Patterson-Stallwood L, Sayfi S, Motilall A, et al. A multimethods randomized trial found that plain language versions improved parents’ understanding of health recommendations. J Clin Epidemiol. 2023;161:8–19.

10. Bujold M, El Sherif R, Bush PL, Johnson-Lafleur J, Doray G, Pluye P. Ecological content validation of the Information Assessment Method for parents (IAM-parent): A mixed methods study. Eval Program Plann. 2018;66:79–88.

11. Pluye P, Granikov V, Bartlett G, Grad RM, Tang DL, Johnson-Lafleur J, et al. Development and content validation of the information assessment method for patients and consumers. JMIR Res Protoc. 2014;3(1):e7.

12. Shoemaker SJ, Wolf MS, Brach C. The Patient Education Materials Assessment Tool (PEMAT) and User’s Guide. (Prepared by Abt Associates, Inc. under Contract No. HHSA290200900012I, TO 4). Rockville, MD: Agency for Healthcare Research and Quality; November 2013. AHRQ Publication No. x14-0002-EF.

13. Harris PA, Taylor R, Minor BL, Elliott V, Fernandez M, O’Neal L, et al. The REDCap consortium: Building an international community of software platform partners. J Biomed Inform. 2019;95:103208.

14. Harris PA, Taylor R, Thielke R, Payne J, Gonzalez N, Conde JG. Research electronic data capture (REDCap)--a metadata-driven methodology and workflow process for providing translational research informatics support. J Biomed Inform. 2009;42(2):377–81.

15. Braun V, Clarke V. Conceptual and design thinking for thematic analysis. Qualitative Psychology. 2022;9(1):3.

16. Virzi RA. Refining the Test Phase of Usability Evaluation: How Many Subjects Is Enough? Human Factors. 1992;34(4):457–68.

17. Légaré F, Stacey D, Forest PG, Coutu MF. Moving SDM forward in Canada: milestones, public involvement, and barriers that remain. Z Evid Fortbild Qual Gesundhwes. 2011;105(4):245–53.

